# Mathematical analysis of the overall survival after chemoradiotherapy of limited-stage small cell lung cancer and the effect of dose/fractionation

**DOI:** 10.64898/2026.06.11.26355440

**Authors:** Arnau Buñuel-Muriscot, Isabel González-Crespo, Pedro Otero-Casal, Antonio Gómez-Caamaño, Juan Pardo-Montero

**Author notes:** **Correspondence:** Juan Pardo-Montero, Grupo de Física Médica e Biomatemáticas, Instituto de Investigación Sanitaria de Santiago (IDIS), Servizo de Radiofísica e Protección Radiolóxica, Hospital Clínico Universitario de Santiago, Trav. Choupana s/n, 15706, Santiago de Compostela (Spain). Now at NOW Systems, Santiago de Compostela, Spain. Now at Institute of Analysis and Scientific Computing, TU Wien, Vienna, Austria.

## Abstract

The purpose of this work is to analyze the 2-year overall survival (OS_2y_) of limited-stage small cell lung cancer (LS-SCLC) treated with chemoradiotherapy (CRT), aiming at characterizing the response of LS-SCLC, and in particular the *α*/*β* value and proliferation parameters. Through a systematic analysis of the literature, we collated a dataset containing 57 entries (3363 patients) of response of LS-SCLC treated with CRT. Radiotherapy schedules ranged from hyper- to hypofractionation. Four radiobiological models to describe the OS_2y_ were investigated, with progressive levels of complexity including the effect of radiotherapy, chemotherapy, treatment year and toxicity. The Akaike Information Criterion (AIC) was used to compare models, and the profile likelihood methodology to compute confidence intervals. Model 4, which includes the effect of radiotherapy, chemotherapy, treatment year and dose-dependent toxicity, provided the best fits of the experimental data (lowest AIC value). While being the best model, model 4 still fails to provide a good prediction of the OS_2y_, in particular failing to predict the survival of the schedules achieving the lower/higher survivals. The radiobiological analysis of the dose-response of LS-SCLC to CRT does not allow to narrowly constrain the value of response parameters. We attribute this limitation to the large heterogeneity of this disease. Nonetheless, our analysis shows a large *α*/*β* value (>9 Gy, 95% CI), which implies a low fractionation effect in the radiotherapy of LS-SCLC. and an accelerated proliferation of tumor cells, *λ*^′^ > 1.6 Gy day^−1^ (95% CI), after a kick-off time of ∼4-5 weeks, which supports the use of accelerated protocols to avoid the effect of tumor proliferation on the clinical outcome.

## 1. Introduction

Limited-stage small cell lung cancer (LS-SCLC) is usually treated with chemoradiotherapy (CRT). Improvements in both radiotherapy (RT) techniques and chemotherapy regimes have increased the overall survival (OS) in the last 20 years, which nonetheless remains low, with typical values ranging from 30% to 60% at two years [1–3].

Treatment advances in the last 20 years have been modest, and include the optimization of radiotherapy and chemotherapy protocols. SCLC exhibits rapid growth, and the acceleration of RT may limit tumor proliferation and improve clinical outcomes. This rationale has led to the use of accelerated two daily fractions radiotherapy protocols to treat LS-SCLC [1]. Also, hypofractionated schedules delivering in some cases doses far larger than 2 Gy/fraction have been used to treat LS-SCLC, including extreme hypofractionation [4]. In very recent times, immune checkpoint inhibitors and novel chemotherapy agents may lead to an improvement of the clinical outcomes for LS-SCLC [1, 5].

Radiobiological modeling has been widely used to analyze the dose-response of many cancers, as it allows to obtain important response parameters (in particular, the *α*/*β* ratio that characterizes the response to fractionation, and the proliferation parameters) that can later assist in the design of optimal RT protocols [6–8]. However, mathematical analyses of the response of LS-SCLC to RT (and CRT) are scarce. A recent study by Li et al. [9] presented a novel mathematical model that analyzed not only the outcome but also the shape of the Kaplan-Meier curve (outcome versus time). When applied to analyze the local progression-free survival of extended-stage SCLC (two schedules delivering 45 Gy/30 fractions/twice-daily and 45 Gy/25 fractions/once-daily), they obtained *α*/*β*=9.2 Gy. A recent study by Diao et al. [10] investigated the fractionation debate in LS-SCLC by fixing *α*/*β*=10 Gy. In this work, we further explored the radiobiology of LS-SCLC treated with CRT with a large dataset of treatments, with doses-per-fraction ranging from 1.2 Gy to 12 Gy. We focused on the analysis of the overall survival at two years, and we built on a recent study by Nix et al. [6], who presented several models to analyze the OS of non-small cell lung cancer (NSCLC) treated with CRT.

## 2. Materials and methods

### 2.1. Clinical dataset

We analyzed the PubMed database to find clinical studies reporting clinical outcomes for the chemoradiotherapy of LS-SCLC. Different search strings were tried to minimize the probability of missing relevant studies, e.g. *“SCLC & radiotherapy & control & trial”*. The literature search was finished in May 2023. We restricted our analysis to overall survival at 2 years, OS_2y_, because we observed that it was the most frequently reported outcome. Kaplan-Meier OS_2y_ were generally reported in the text but sometimes were extracted from figures by using the image analysis software g3data. In addition to OS_2y_, we also extracted information regarding total dose (*D*), dose per fraction (*d*), treatment time (*T*), number of patients in the study (*N*), year of the study, and concurrent or sequential chemotherapy. The studies were discarded if any of this information was missing. In addition, we extracted the following complementary information when available: age (median), sex (% of male patients), use of prophylactic cranial irradiation, PCI (% of patients), chemotherapy agents, T stage, N stage, AJCC stage and performance score, PS. These extra variables were not included in the mechanistic models, but were included in a multivariable meta-regression analysis.

We limited our analysis to studies published post-1995 (included). Overall, the collated dataset contained 57 entries (3363 patients) obtained from 39 publications [4,11–48]. The full dataset is presented in Supplementary Table 1.

### 2.2. Radiobiological modeling

We used the linear-quadratic (LQ) model to describe the effect of radiotherapy. As several schedules deliver multiple fractions per day, we used the LQ with incomplete repair correction [49,50]. The surviving fraction of cells following the i-th radiation fraction is given by:

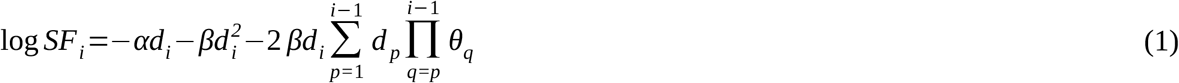

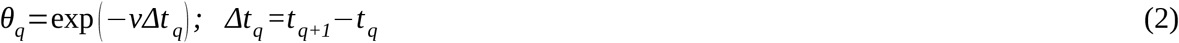

where *α* and *β* are the LQ parameters characterizing cell radiosensitivity, and *ν* is the repair rate of sublethal damage (the half-life of damage is defined as *T*_*repair*_ = log 2/*ν*).

For a treatment delivering n fractions, the overall surviving fraction is,

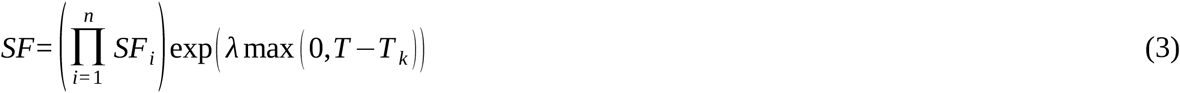

where *SF*_*i*_ is the surviving fraction of fraction *i, T* is the overall treatment time, and accelerated proliferation is modeled as an exponential function with proliferation rate *λ* after a kick-off time *T*_*k*_.

Starting from the LQ model, four different models were used to fit the OS_2y_ data based on the models presented in Nix et al. [6] to model OS for advanced NSCLC.

#### Model 1: logistic function for the RT effect

The simplest model, model 1, uses a logistic function for the overall survival, similar to logistic models used to describe tumor control probability after radiotherapy:

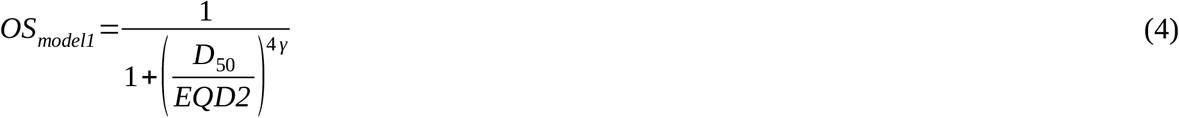

where *D*_*50*_ is the dose corresponding to 50% survival (in 2 Gy fractions) and *γ* is the normalised dose-response gradient. *EQD2* is the equivalent dose in 2 Gy fractions of a given schedule, which assuming that every fraction delivers the same dose is calculated as:

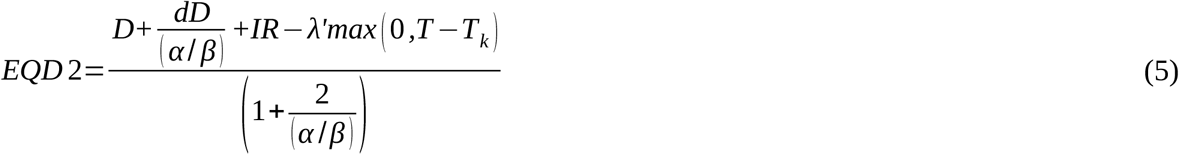

In this equation, *λ*^′^ = *λ*/*α*, and *IR* denotes the effect of incomplete repair on the EQD2. This term can be written as:

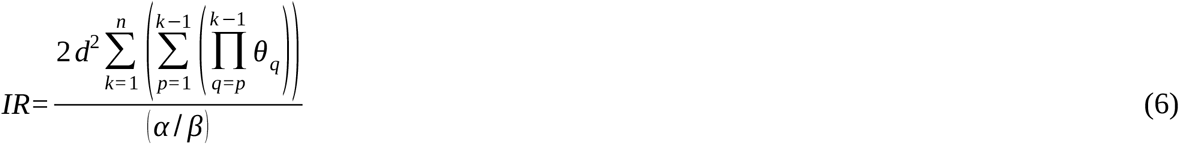

#### Model 2: concurrent/sequential CT effect

Model 2 builds on model 1 and specifically incorporates the effect of chemotherapy as:

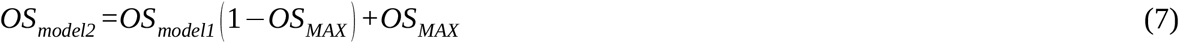

where *OS*_*MAX*_ is a fitting parameter which accounts for the effect of chemotherapy. The rationale behind this model is that chemotherapy alone leads to the survival of a fraction *OS*_*MAX*_ of the patients, and the addition of radiotherapy will increase the survival by affecting the fraction (1-*OS*_*MAX*_). In addition, because there is evidence of better results when delivering concurrent rather than sequential chemoradiotherapy, the *EQD*2 of those schedules was scaled up as:

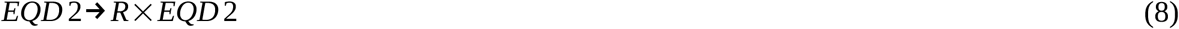

#### Model 3: inclusion of year of treatment

We observed a positive correlation between the publication year and survival in the pre-analysis of the dataset, which can be caused by progressive improvements in the chemotherapy and radiotherapy protocols over the years (a similar pattern was observed in [6] for NSCLC). Therefore, we used the following model:

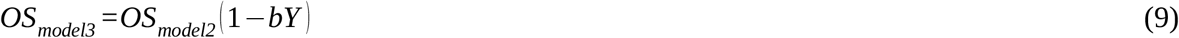

where *b* is a fitting parameter and *Y*=2023-publication year.

#### Model 4: logistic function for RT toxicity

The fourth model accounts for the possibility of decreasing survival with increasing radiation dose due to increased radiation toxicity, and models OS_2y_ as:

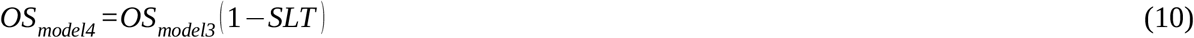

where *SLT* (survival limiting toxicity) is a logistic function like in Equation (1),

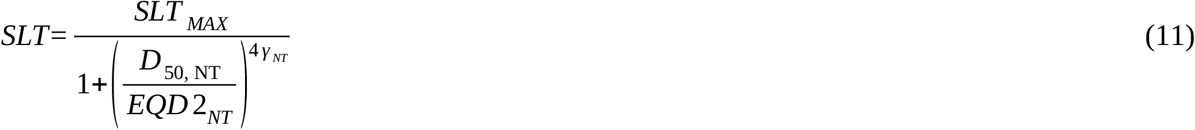

and the equivalent dose in 2 Gy fractions is calculated ignoring proliferation and sublethal damage repair, and for a fixed *α*/*β*=3 Gy.

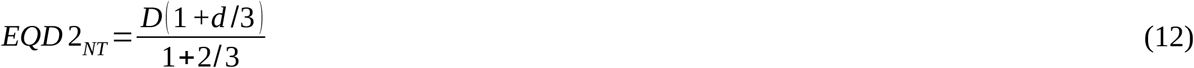

#### 2.3. Statistical methods and implementation

A preliminary multivariable meta-regression using inverse variance weighting was performed to identify factors influencing the OS_2y_. We used the maximum likelihood methodology to fit the models to the clinical data, assuming binomial statistics for the reported OS. The optimization (minimization of *-log L*, where *L* is the likelihood) was performed with an in-house developed algorithm based on the simulated annealing method [8]. The fitting parameters were *α*/*β, λ*^′^, *T*_*repair*_, *T*_*k*_, *γ*, and *D*_*50*_ for Model 1, with two extra fitting parameters for Model 2 (*OS*_*MAX*_ and *R*), an extra parameter for Model 3 (*b*), and two extra parameters for Model 4 (*γ*_NT_, and *D*_*50,NT*_). After an initial optimization, outliers were identified (they were defined as studies i for which the probability *L*_*i*_ of the experimental OS matching the predicted OS was below 10^−5^) and a second optimization was performed excluding those outliers to investigate potential biases in the best-fitting parameters caused by them.

The searchable space for parameter values was constrained to avoid reaching solutions that could be unphysical or not supported by biological data, and to speed up convergence. In particular, we employed the maximum and minimum bounds reported in Table 1. In order to obtain confidence intervals (CI) for the best-fitting parameters we used the profile likelihood method. The chi-square test was used to evaluate the goodness of fit by computing *p* values and χ^2^/dof. The Akaike Information Criterion with sample size correction (AIC_c_) was used to evaluate the performance of the models.

**Table 1.**
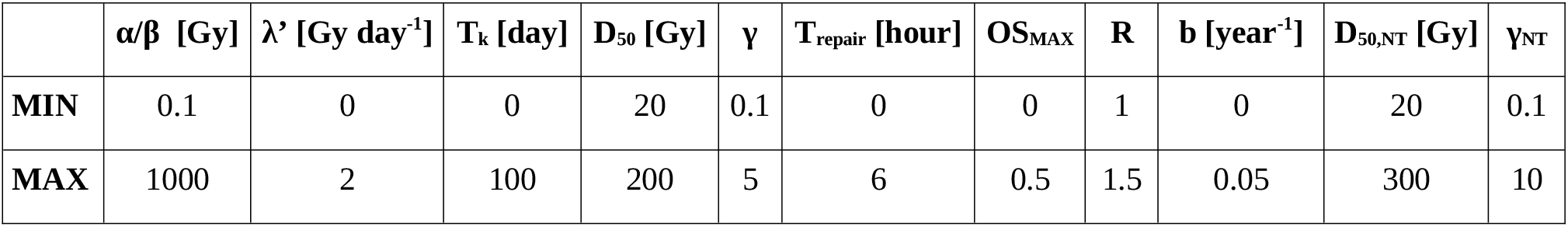
Minimum (MIN) and maximum (MAX) constraints on the best-fitting parameter values.

An approximation was used in the computation of the sublethal damage repair to speed up the calculations: when computing the sublethal damage repair contribution for a fraction *i*, only fraction *i-1* was considered, rather than all fractions from 1 to *i-1*; the rationale for this is that the rate of sublethal damage repair was limited to *T*_*repair*_<6 h and no schedules deliver more than 2 fractions per day, therefore the time between fractions *i-2* and *i* will be >24h (>4 half-life); this approximation speeded the calculation up to a factor 10.

The implementation of the methodology was performed in Matlab (The Mathworks, Natick, MA).

## 3. Results

In a multivariable meta-regression including overall treatment time, dose, dose per fraction, study year, concurrent or sequential chemotherapy, age, sex, PCI, and PS, only the dose per fraction (*p*<0.01), total dose (*p*<0.02) and sex (*p*=0.03) were significantly correlated with OS_2y_. However, because the dataset missed many entries for some of these variables, the study was limited to 42 schedules out of 57.

A second multivariable meta-regression was performed limited to overall treatment time, dose, dose-per-fraction, treatment year, and concurrent/sequential chemotherapy, matching the variables used in the radiobiological models. The results are shown in Table 2. In this case, the most significant predictor is the study year (this is illustrated in Figure 1), followed by total dose, treatment time, and dose per fraction.

**Table 2.** Results of a multivariable meta-regression using inverse variance weighting and including the variables overall treatment time, total dose, dose per fraction, study year, and concurrent/sequential chemotherapy.

| Variable | regression coefficient | p value |
| --- | --- | --- |
| dose per fraction | -0.039 | 0.074 |
| total dose | 0.007 | 0.040 |
| treatment time | -0.006 | 0.069 |
| study year | 0.006 | 0.008 |
| sCRT | -0.010 | 0.853 |

**Figure 1.**
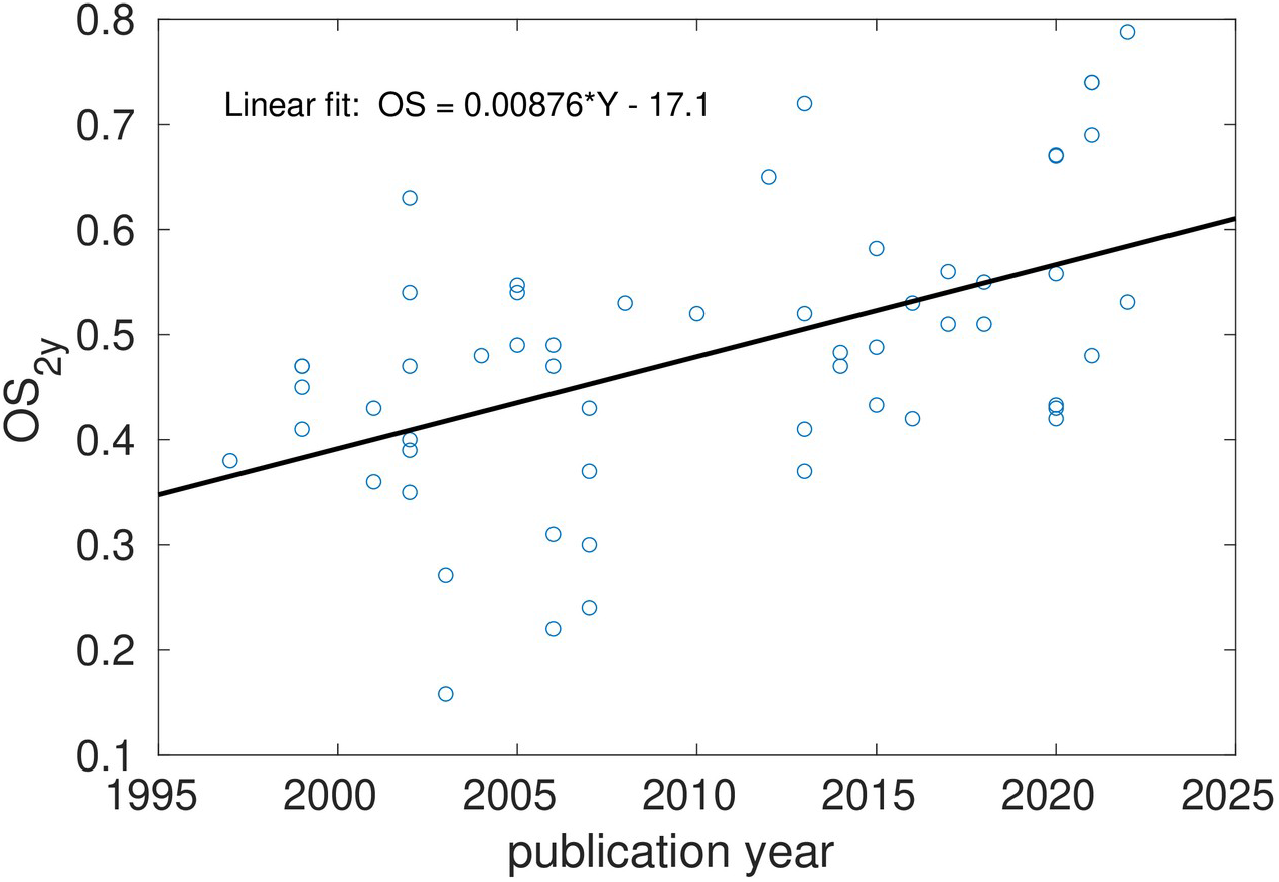
Overall survival at two years versus publication year of the study: clinical points and linear fit.

The initial optimizations of Models 1-4 systematically showed two outliers (indices 41 and 48 in Supplementary Table 1). These outliers had very low OS_2y_ (0.22 and 0.16, respectively), which none of the models was able to reproduce. The optimizations were re-run after removing those outliers. In Table 3 we report best-fitting parameters for each of the four models used in this work, together with goodness-of-fit (-*log L*) and *AIC*_*c*_ values. All models lead to best fits showing large *α*/*β* values and important proliferation after ∼5 weeks. The removal of the outliers did not lead to very different best-fitting parameters (results not shown).

**Table 3.**
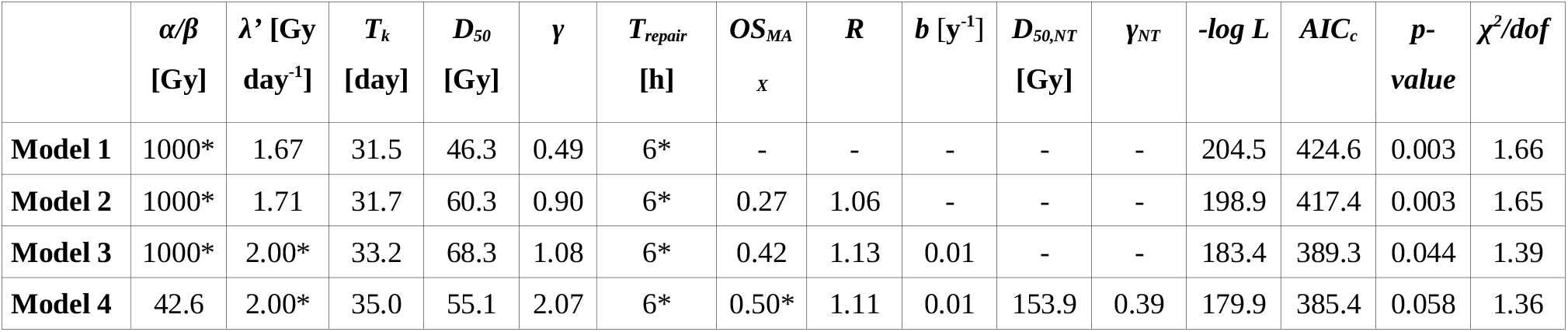
Best-fitting parameters, value of the *-log L* objective function, Akaike Information Criterion value (*AIC*_*c*_), goodness of fit (evaluated with *p* values and *χ*^*2*^*/dof*) for each model. The symbol * indicates that a parameter has reached the maximum or minimum bound of the space of values allowed for that particular parameter.

Models 3 and 4 provided better fits, with differences in AIC_c_ around 30–40 when compared with Models 1 and 2. Because of this, the 95% confidence interval calculation for the best-fitting parameters was limited to models 3 and 4. Those intervals are reported in Table 4.

**Table 4.** 95% confidence intervals of the best-fitting parameters for model 3 and model 4, obtained through the profile likelihood methodology.

|  | **Model 3** | **Model 4** |
| --- | --- | --- |
| $\alpha/\beta$ [Gy] | [30.6, 1000] | [8.9, 1000] |
| $\lambda'$ [Gy day<sup>-1</sup>] | [1.6, 2.0] | [1.6, 2.0] |
| $T_k$ [day] | [30.5, 35.6] | [32.1, 36.7] |
| $D_{50}$ [Gy] | [56.1, 94.2] | [48.2, 68.7] |
| $\gamma$ | [0.54, 1.89] | [1.04, 3.37] |
| $T_{repair}$ [h] | [0, 6] | [0, 6] |
| $OS_{MAX}$ | [0.22, 0.50] | [0.35, 0.50] |
| $R$ | [1.00, 1.47] | [1.01, 1.25] |
| $b$ [y<sup>-1</sup>] | $[6.5, 12.7] \times 10^{-3}$ | $[6.5, 12.9] \times 10^{-3}$ |
| $D_{50,NT}$ [Gy] | - | [102.8, 1000] |
| $\gamma_{NT}$ | - | [0.13, 0.66] |

In Figure 2 we report the model-predicted OS_2y_ for each schedule as well as the 95% confidence intervals of the experimental data (calculated by assuming a binomial distribution). The best of the models, Model 4, fails to predict an OS_2y_ within the 95% confidence interval in 9/55 schedules, a 17% of the schedules, above the statistical 5% expectation.

**Figure 2.**
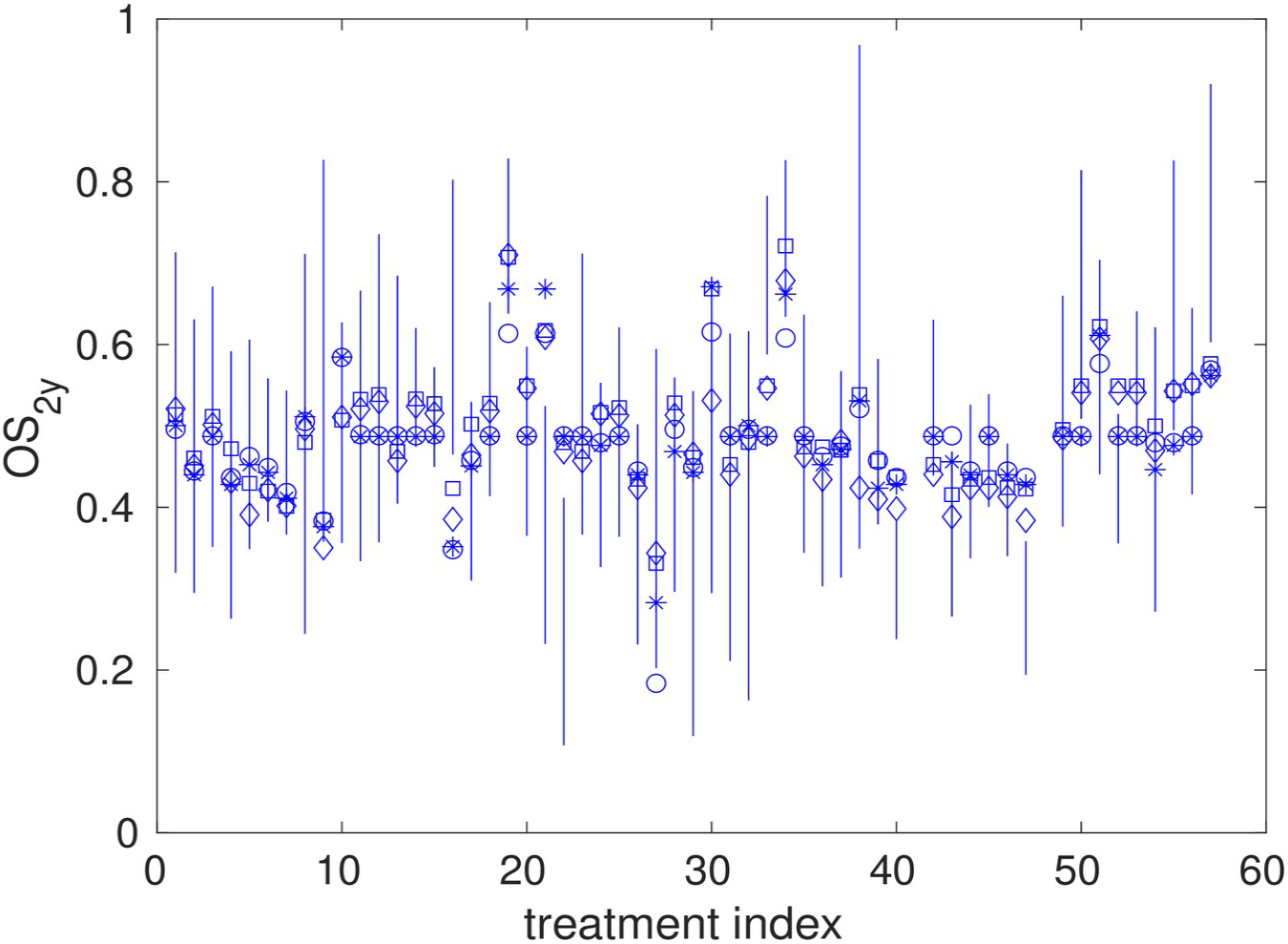
Binomial 95% confidence intervals for the overall survival at 2 years (OS_2y_) for each trial analyzed in this study (solid lines), and model predictions (Model 1, circles; Model 2, asterisks; Model 3, squares; Model 4, diamonds).

It can be qualitatively noticed in Figure 2 that the models fail to predict the OS_2y_ of schedules with low and large survival rates, overestimating the survival of the former and underestimating the survival of the latter. This is more easily noticed in Figure 3, where the clinical and model predicted sorted OS_2y_ are plotted.

**Figure 3.**
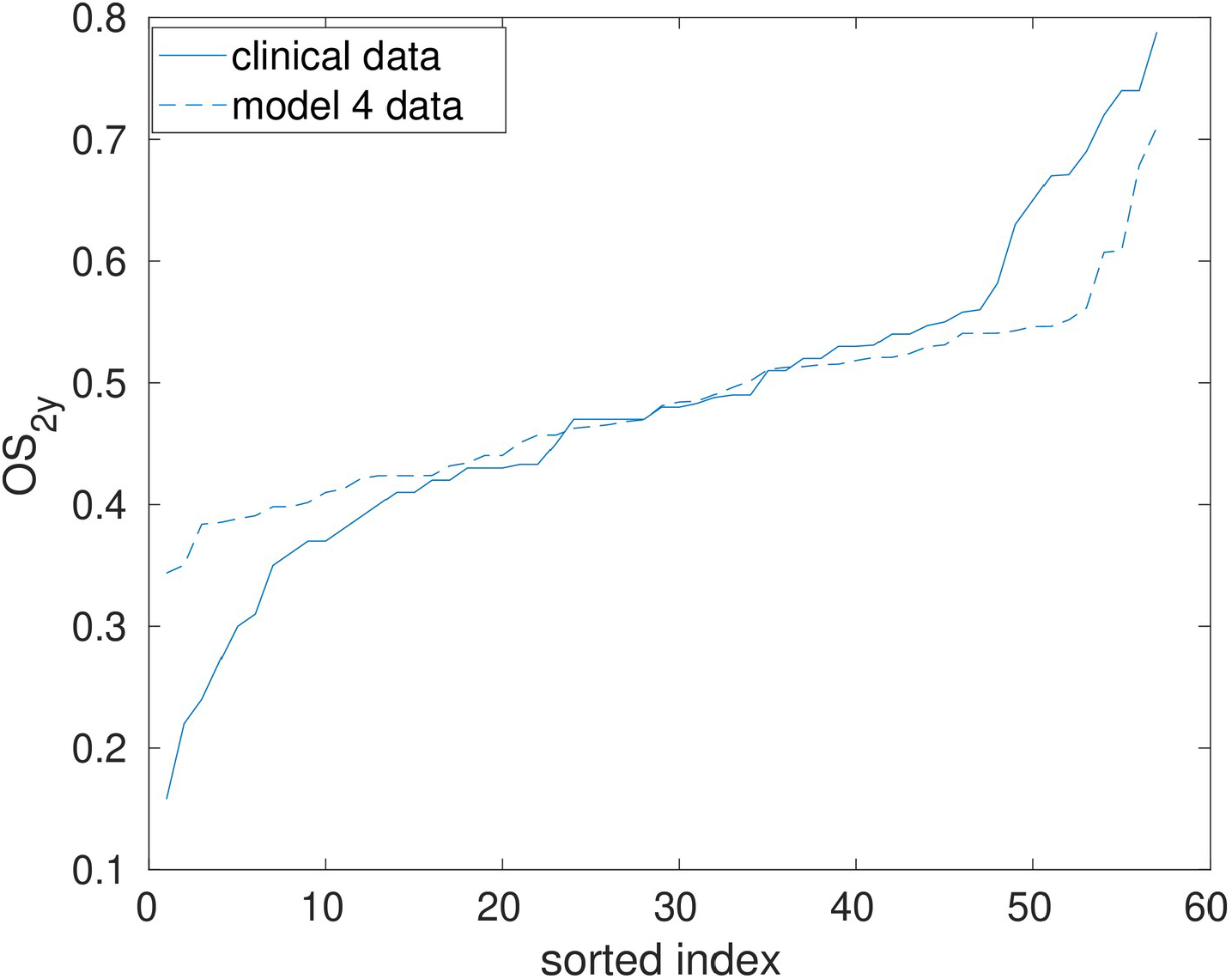
Sorted values of the overall survival at 2 years (OS_2y_) for each trial analyzed in this study: clinical values (solid line), and model 4 predictions (dashed line). The model fails to predict the survival of schedules presenting low/large OS_2y_ values.

## 4. Discussion

Limited-stage small cell lung cancer is usually treated with chemoradiotherapy. The radiotherapy schedules used to treat LS-SCLC are diverse and include conventional 2 Gy/fraction schedules, accelerated protocols delivering multiple fractions per day, and hypofractionation. Acceleration and hypofractionation may limit the effect of tumor proliferation and improve clinical outcomes.

The radiobiological modeling of the cancer dose-response can provide important response parameters, in particular, the *α*/*β* ratio that characterizes the response to fractionation, and the proliferation parameters that characterize the diminishing effectiveness with increasing treatment duration. The determination of those parameters can later assist in the design of optimal RT protocols [6–8]. This type of analysis has been conducted for many cancers, but the mathematical analyses of the response of LS-SCLC to CRT are scarce.

In this work, we have analyzed the dose-response of LS-SCLC to CRT. To this aim, we have collated a dataset of 57 entries (3363 patients), and analyzed the overall survival at 2 years with four dose-response models with increasing complexity. Those models are based on a previous study by Nix et al. [6] who investigated the overall survival of NSCLC treated with CRT.

Models 1 and 2, which include the effect of radiotherapy on the tumor and chemotherapy, failed to fit the collated dataset as shown by the *p* values and χ^2^/dof of the chi-square test (*p ∼* 0.003, χ^2^/dof ∼ 1.6). Models 3 and 4, which add the effect of the study year and the dose-induced toxicity, clearly improve upon Model 1 and 2, with Model 4 reaching *p=*0.06, χ^2^/dof=1.36.

In this dataset we did not correct for follow-up. Patients lost to follow-up may result in an *effective* number of patients below the reported number of patients in the trial for statistics computation and therefore an underestimation of the uncertainties [51, 52]. This may contribute to the observed *p* and χ^2^/dof values: as an illustration, assuming that 10% of the patients of each study were lost to follow-up, Model 4 yields *p=*0.15, χ^2^/dof=1.22, and 20% of patients lost to follow-up leads to *p=*0.32, χ^2^/dof=1.09.

The AIC analysis clearly confirmed Model 4 to be the best of the models (lowest *AIC*_*c*_ value), with differences in AIC reaching up to 40 compared to Model 1. While being the best model, model 4 still fails to provide a good prediction of the OS_2y_, in particular failing to predict the survival of the schedules achieving the lower/higher survivals.

LS-SCLC is a heterogeneous disease treated with various protocols. In particular, the different studies included in the dataset may use different margins, tissue heterogeneity correction algorithms, chemotherapy drugs, or treat populations of patients with different characteristics. This increases the heterogeneity of the dataset and therefore may result in increasing fitting uncertainties.

The radiobiological analysis of the dose-response of LS-SCLC to CRT performed in this work does not allow to narrowly constrain the value of response parameters. Nonetheless, our analysis is consistent with a large *α*/*β* value (>9 Gy, 95% CI), which implies a low fractionation effect in the RT of LS-SCLC. Also, our analysis is consistent with an accelerated proliferation of tumor cells, *λ*^*’*^ >1.6 Gy day^−1^, after a kick-off time of ∼4–5 weeks. For accelerated radiotherapy delivering multiple fractions per day, the quantification of the rate of sublethal damage repair is important to evaluate the possible incomplete repair contribution. However, this analysis was not sensitive to changes in *T*_*repair*_ (see 95% CIs reported in Table 4). Altogether, this analysis supports the use of accelerated or hypofractionated protocols to avoid the effect of tumor proliferation on the clinical outcome. Also, both Model 4 and the multivariable meta-regression are consistent with a decrease in OS when the dose/dose-per-fraction is large, perhaps due to dose-induced toxicity.

It is important to acknowledge that the clinical landscape of LS-SCLC has undergone a fundamental shift in very recent years with the emergence of immunotherapy (IT) drugs which have significantly increased overall survival [53]. This may trigger the study of new models including the addition of IT. More advanced models may also provide a better description of the treatment outcomes. In particular, the modeling of the effect of chemotherapy that we have used in this work is rather simple, and it could be improved by considering more realistic models that for example take into account differences in the chemotherapy drugs and the number of cycles [54]. In this regard, the collated dataset could be useful to develop more advanced models and to obtain a better characterization of the dose-response of LS-SCLC to CRT.

## Data Availability

All data produced in the present work are contained in the manuscript.

## Acknowledgements

This project has received funding from Ministerio de Ciencia e Innovación, Agencia Estatal de Investigación and FEDER, UE (grant PID2021-128984OB-I00), and Xunta de Galicia, Axencia Galega de Innovación (grant IN607D 2022/02).

## Data Availability Statement

All data used in this work are available in the article and the supplementary materials.

## Supplementary Materials

**Table S1:**
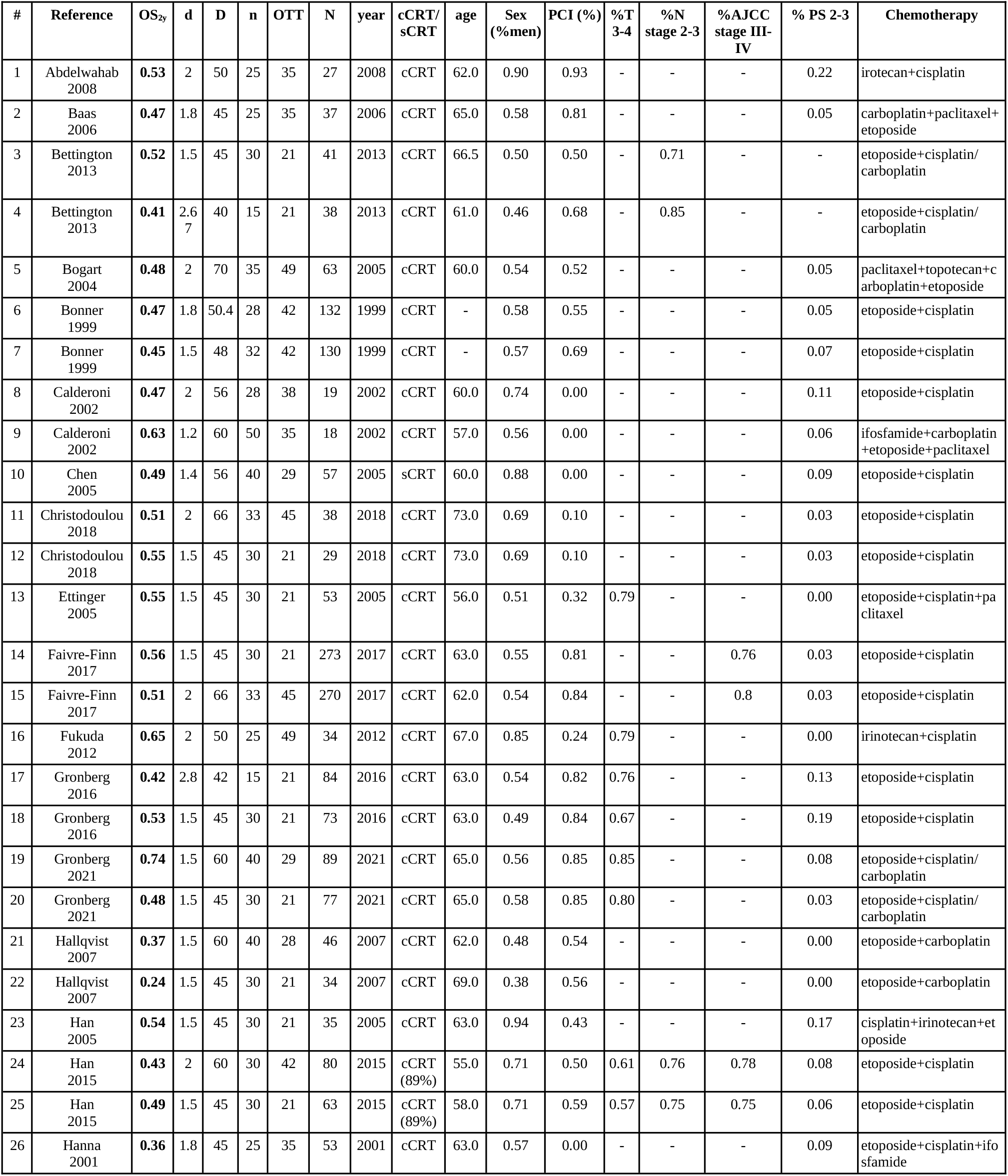
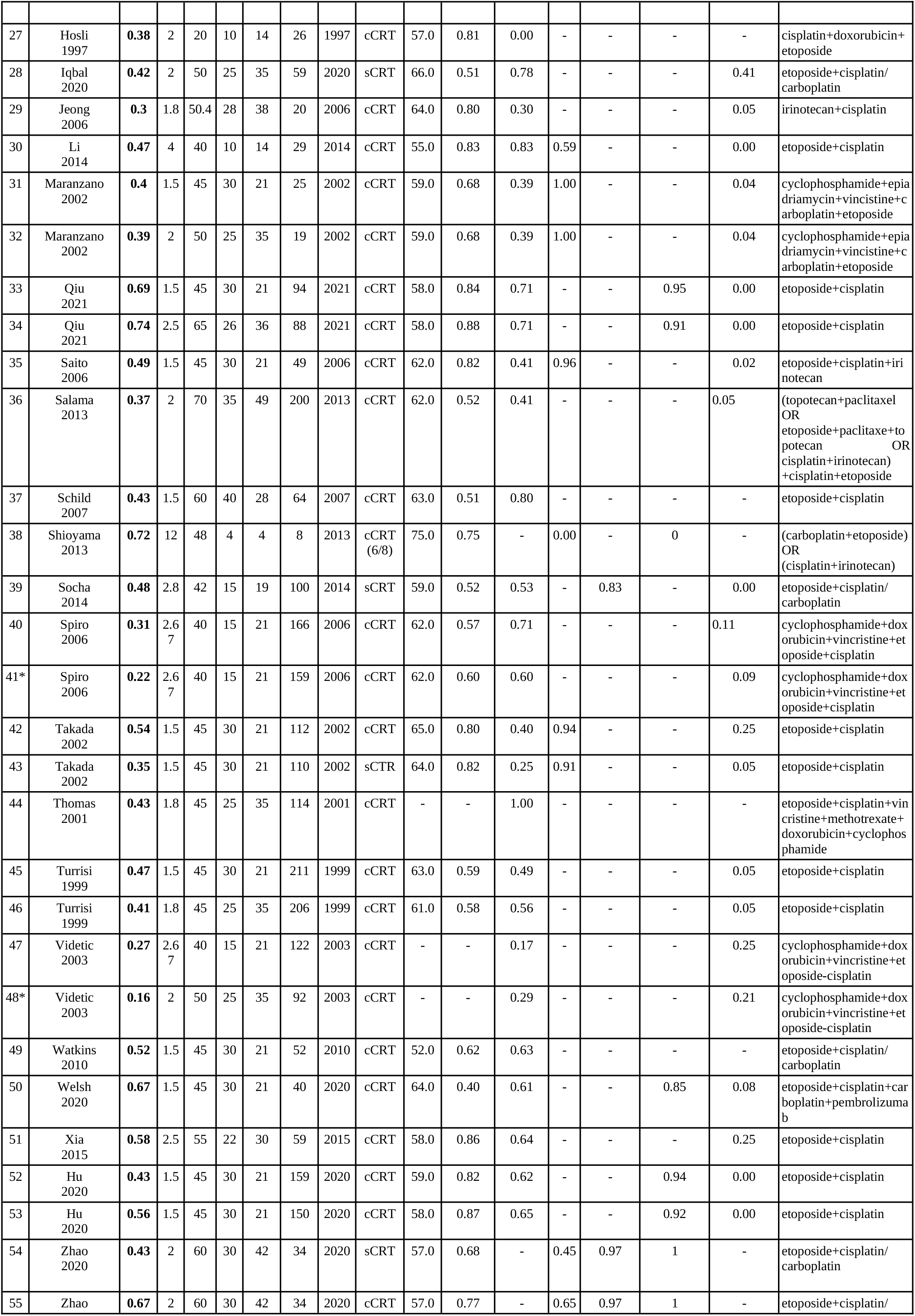
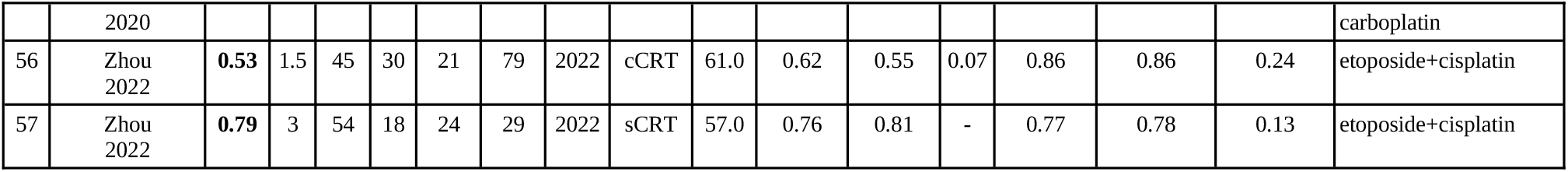
Dataset collated for this study. For each entry we report overall survival at 2 years (OS_2y_), dose per fraction (d), total dose (D), number of fractions (n), radiotherapy overall treatment time (OTT), number of patients (N), treatment year (year), and conformal/sequential chemoradiotherapy (cCRT/sCRT), and chemotherapy drugs. When available, we also report when available median age, sex (fraction of male patients), fraction of patients receiving prophylactic cranial irradiation (PCI), fraction of patients with T-stage 2-3, N-stage 2-3, AJCC stage 3-4, and performance status (PS) 2-3. The entries #41 and #48 (marked with an asterisk) were considered outliers and removed from the analysis.

